# Model-based Evaluation of Continued COVID-19 Risk at Long Term Care Facilities

**DOI:** 10.1101/2021.07.06.21259931

**Authors:** Bailey K. Fosdick, Jude Bayham, Jake Dilliott, Gregory D. Ebel, Nicole Ehrhart

**Affiliations:** Department of Statistics, Colorado State University, Ft. Collins, CO 80523; Department of Agricultural and Resource Economics, Colorado State University, Ft. Collins, CO 80523; Arthropod-Borne and Infectious Diseases Laboratory, Department of Microbiology, Immunology and Pathology, Colorado State University, Ft. Collins, CO 80526; Columbine Health Systems Center for Healthy Aging and Department of Clinical Sciences, Colorado State University, Fort Collins, CO 80523

**Keywords:** agent-based model, nonpharmaceutical interventions, vaccine uptake, COVID-19

## Abstract

The COVID-19 pandemic severely impacted long-term care facilities resulting in the death of approximately 8% of residents nationwide. As COVID-19 case rates decline and state and county restrictions are lifted, facility managers, local and state health agencies are challenged with defining their own policies moving forward to appropriately mitigate disease transmission. The continued emergence of variants of concern has highlighted the need for a readily available tool that can be employed at the facility-level to determine best practices for mitigation and ensure resident and staff safety. To assist leadership in determining the impact of various infection surveillance and response strategies, we developed an agent-based model and an online dashboard interface that simulates COVID-19 infection within congregate care settings under various mitigation measures. In this paper, we demonstrate how this dashboard can be used to quantify the continued risk for COVID-19 infections within a facility given a designated testing schedule and vaccine requirements. Our results highlight the critical nature of testing cadence, test sensitivity and specificity, and the impact of removing asymptomatic infected individuals from the workplace. We also show that monthly surveillance testing at long-term care facilities is unlikely to successfully mitigate SARS-CoV-2 outbreaks in congregate care settings.

**Disclosures:** This work was supported by Colorado State University’s Center for Healthy Aging, the Center for Vector-Bourne Infectious Disease, the Office of the Vice President for Research, the College of Health and Human Sciences, the Collage of Natural Sciences, the College of Veterinary Medicine and Biomedical Sciences, and the Walter Scott Jr College of Engineering.

## Introduction

Nursing homes, skilled nursing facilities and assisted living facilities, collectively known as long-term care facilities (LTCFs), provide care for some of the most vulnerable populations in society. Shared sleeping quarters, bathrooms, dining facilities, and common spaces and the need for daily contact between staff and residents create an opportunistic environment for the spread of any infectious disease. In particular, the morbidity and mortality within LTCFs throughout the COVID-19 pandemic is estimated at nearly 8% and has demonstrated the extreme vulnerability of both LTCF residents and staff to a transmissible viral illness.^1^ Approximately 40% of COVID-related deaths in the United States have been among residents of LTCFs,^2^ and yet clinical research in LTCFs during COVID-19 has been limited.^3^ Moreover, LTCF staff represent a disproportionately high percentage of SARS-CoV-2 infections as compared to non-healthcare community members regardless of whether or not LTCF staff have direct patient contact. In one report, whole-genome sequencing suggested that SARS-CoV-2 infection among LTCF staff had more likely come from staff-staff transmission than community import events.^4^

In recognition of the difficulty mitigating or preventing SARS-CoV-2 within LTCFs, the Centers for Disease Control (CDC) recommended that LTCFs have first priority for vaccine access and developed The Pharmacy Partnership for Long Term Care Program to distribute the first available vaccines to LTCF residents and staff. However, early data suggest that there has been low vaccine acceptance among LTCF staff. A recent study reported that among the 11,460 LTCFs with at least one vaccination clinic conducted via the CDC Pharmacy Partnership for Long Term Care Program, only 37.5% of staff members received the vaccine as compared to 77.8% of residents.^5^ These data are concerning because unvaccinated staff can sustain SARS CoV-2 infection within LTCFs, making infection control extremely difficult, even with higher vaccination rates, new variants (e.g., delta variant) pose a risk because they may be more resistant to the vaccine.^6,7^ In a study of infections in LTCFs in Catalonia, Spain during spring 2021, researchers found that once more than 70% of the facility population was vaccinated, approximately 75% of COVID-19 deaths and infections in the facility were prevented.^8^ Vaccination rates are of critical importance even for LTCF residents that were previously infected with COVID-19 as a substantial proportion have been found to have nondectable antibodies six months post infection.^9^

Centers for Medicare and Medicaid Services (CMS) state and local public health recommendations for SARS-CoV-2 surveillance testing in LTCFs to identify and isolate presymptomatic/ asymptomatic SARS-CoV-2 positive individuals have evolved over time. A number of learnings have emerged throughout the pandemic that highlight limitations with a “one size fits all” approach to surveillance and outbreak response in LTCF’s. Mitigation and prevention approaches rarely consider the simultaneous influences of test type, predicted sensitivity/specificity, testing frequency, testing goal (surveillance versus diagnostic), test result latency period to inform testing guidance and vaccine acceptance rates. Given the rapidly changing climate surrounding COVID-19 prevalence, testing availability, vaccination acceptance, and community prevalence, we sought to create a model to better understand potential outcomes within LTCFs using inputs related to these real-life and fluid variables as they change throughout time.

Agent-based models (ABMs) are a powerful tool for understanding complex dynamic processes, such as infectious disease transmission.^10^ During the COVID-19 pandemic, these models have been used to assess the impact of nonpharmaceutical interventions on infections within schools^11^, a small town^12^, and France.^13^ Researchers further demonstrated that delaying the second dose of mRNA vaccines for those under 65 led to more positive outcomes under certain conditions with an ABM^14^. Within LTCFs specifically, ABMs were used to study the impact of testing and immunity-based staffing interventions^15^, and early in the pandemic were used to simulate the spatiotemporal transmission process of COVID-19 under varied virus infectiousness levels and within-facility mobility restrictions.^16^ An advantage of ABMs over compartmental models based on differential equations is that they can be programmed to capture important micro-level behavior unique to a specific setting. The agent-based framework has several advantages for modeling LTCFs. First, LTCFs are focused settings with relatively low populations. Second, the ABM permits simple tracking of individuals’ health and behavior over time. This point is important in the LTCF setting, where staff are transient members of the population as they move in and out of the population according to work schedules. Accounting for this behavior is key as infectious individuals only pose a risk to the facility when present. Moreover, ABMs track the state of individual agents in the model. If a member of the night staff is infected, that specific agent can be isolated until they no longer pose an infection risk to other agents in the facility. These features make the ABM a suitable framework for a decision support tool used to analyze the impact of different testing protocols, vaccination rates, and visitation policies.

State and county COVID-19 regulations began to relax heavily in May 2021. This left LTCF managers and local health officials to define their own COVID-19 policies. Some facilities require their staff to be vaccinated, leaning on existing requirements for the influenza vaccine to argue this was a natural policy.^17^ Other facilities have decided not to require vaccination and instead are utilizing surveillance testing to continue to monitor their facility for infection in unvaccinated individuals. While both strategies have advantages and disadvantages, it is unclear how much continued risk a facility is at given a specific level of staff vaccine uptake, testing regime, and community prevalence.

Due to slight nuances across facilities that can greatly affect transmission patterns, we created an interactive online dashboard that accepts facility-specific parameters and forecasts infection rates and worker days missed under a set of transmission characteristics and testing protocols (dashboard is accessible here: https://ltcf-covid.shinyapps.io/ltcf_covid_dashboard/).^18^ This dashboard allows facility administrators to evaluate the relative impact of various strategies moving forward. In what follows, we investigate the trade-off between a high cadence testing regime at a moderately vaccinated facility to a no testing regime at a highly vaccinated facility to demonstrate how the ABM tool might be used in practice.

## Simulation set-up

Consider the scenario where a facility manager is tasked with deciding on policies for summer 2021. The facility has 90 residents, 125 day staff, and 45 night staff. Assume residents and staff are vaccinated at rates mirroring the national averages, 78%, and 38%, respectively, and the 7-day case rate average for the community is approximately 84 people per 100,000/week. The manager is concerned about potential future outbreaks due to their impact on resident and staff physical health, the number of worker days missed due to quarantined staff, and the emotional toll of a return to a lock-down state. Thus, the manager considers a facility-wide vaccination requirement for staff and asks: How many fewer worker days would be missed? How many fewer individuals would be infected with COVID-19? Could infection levels instead be limited by regular testing of unvaccinated staff and residents?

To answer these concrete questions, we program an agent-based model that simulates the key daily behaviors and events that impact disease transmission in a facility. Like the traditional infectious disease compartmental models, all agents (staff and residents) are labeled as either susceptible, exposed, infected, or immune (also referred to as recovered). Each agent transitions between these states based on events in the simulation. For example, all unvaccinated individuals start out as a ‘susceptible’, but after they come into contact with an infectious agent, there is a chance that they contract the virus and become ‘exposed’ for a short period of time before being labeled as ‘infected’ and able to spread COVID-19 to others. Finally, after the active infection period has subsided, the agent moves to the ‘immune’ state. All vaccinated individuals are also given the ‘immune’ label.

The progression of events in a day is described in Figure 1(a). A random fifth of the day staff and fifth of the night staff are selected to show up to work. Each infected staff member has a chance of showing symptoms, self-isolating, and staying home from work. All other staff attend work and have interactions with each other and the residents. While interactions are assumed to be random each day, a contact matrix dictates the relative number of contacts between different subgroups. For example, day staff and night staff are assumed to have limited contact that occurs just during shift changes. Finally, we model staff infection via community transmission based on the local 7-day case rate. Community case rates were shown to be the strongest predictor of COVID-19 cases and outbreaks at LTCFs.^19^ Figure 1(b) depicts the transmission network for the ABM. We assume a vaccine efficacy of 95% in an “all-or-nothing” framework, such that 5% of those vaccinated remain susceptible to infection.^20^

**Figure 1:**
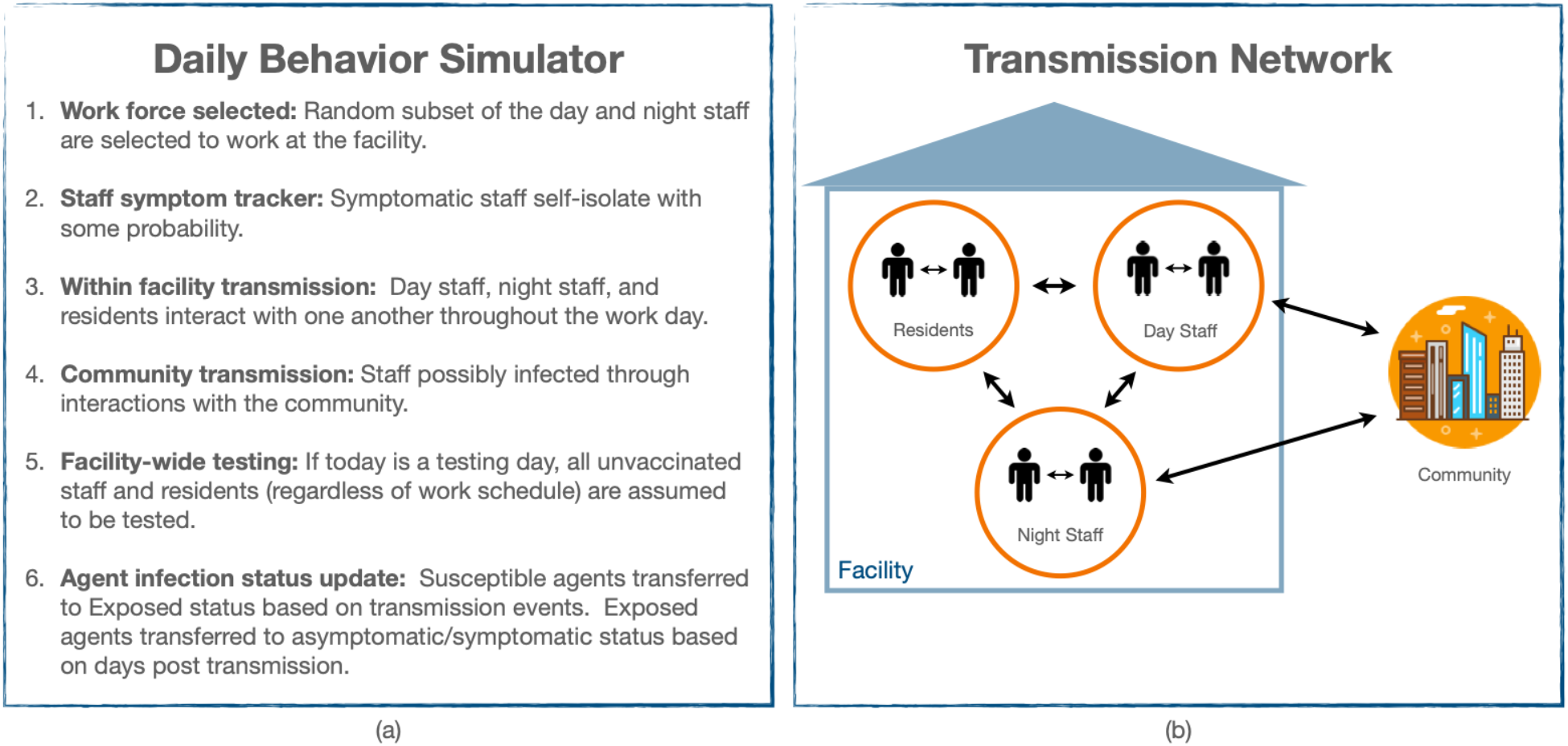
(a) Description of events throughout a day during the simulation. (b) Avenues of transmission within and into the facility.

The primary means for COVID-19 monitoring and outbreak response during the latter half of 2020 and today is facility-wide testing. There are two primary options for surveillance testing: rapid antigen and qRT-PCR tests. The advantage of qRT-PCR tests is that they are extremely accurate, with a sensitivity and specificity greater than 95%; however, the time between the test and receiving the result is often 36-72 hours depending on the lab processing it. Conversely, antigen test results can be acquired the same day but are limited by lower sensitivity. In this study, we assume an qRT-PCR test with 95% sensitivity and 99.5% specificity, and a rapid antigen test with 64.2% sensitivity on symptomatic positive individuals, 35.8% sensitivity on asymptomatic individuals and 99.5% specificity. If vaccine uptake among staff and residents is limited or moderate, a facility director may consider regular facility-wide testing of unvaccinated individuals to combat the possibility and extent of an outbreak. Upon a positive test result, we use a 14-day quarantine for staff.

### Vaccine versus Testing Tradeoff

We simulate the LTCF-ABM to illustrate two continued risks to LTCFs: 1) low staff vaccination rates, and 2) relaxed screening protocols. LTCF staff are a conduit for risk to LTCF residence because they interact with the outside community when not at work. Our model suggests that outbreaks are likely to occur when 78% of residents and 38% of staff are vaccinated even with a weekly testing protocol in place. Figure 2 shows the distribution of the total number of infections across three vaccination scenarios (37.5%, 80%, and 90%) and three testing scenarios (None, qRT-PCR, and Rapid) for 500 simulations. The variation in results across simulations is due to the stochastic transmission functions in the model. The results indicate that while a testing protocol makes a difference, staff vaccination rates are the primary determinant of the total number of infections in both staff and residents. In staff, increasing vaccination from 37.5% to 98% reduces the attack rate by 90% (from 63% to 6%) without a testing protocol. A weekly qRT-PCR-based testing protocol can provide further protection reducing the staff attack rate to a mean of 3% in the 98% staff vaccination case. The combination of high staff vaccination (98%) and weekly qRT-PCR-based testing protocol reduces the attack rate in residents by 82% (from 23% to 4%). In the low staff vaccination scenario, the qRT-PCR-based testing protocol reduces the attack rate by only 11% (from 26% to 23%). The reduced sensitivity of the rapid antigen test has a much smaller effect, especially with relatively low staff vaccination rates.

**Figure 2.**
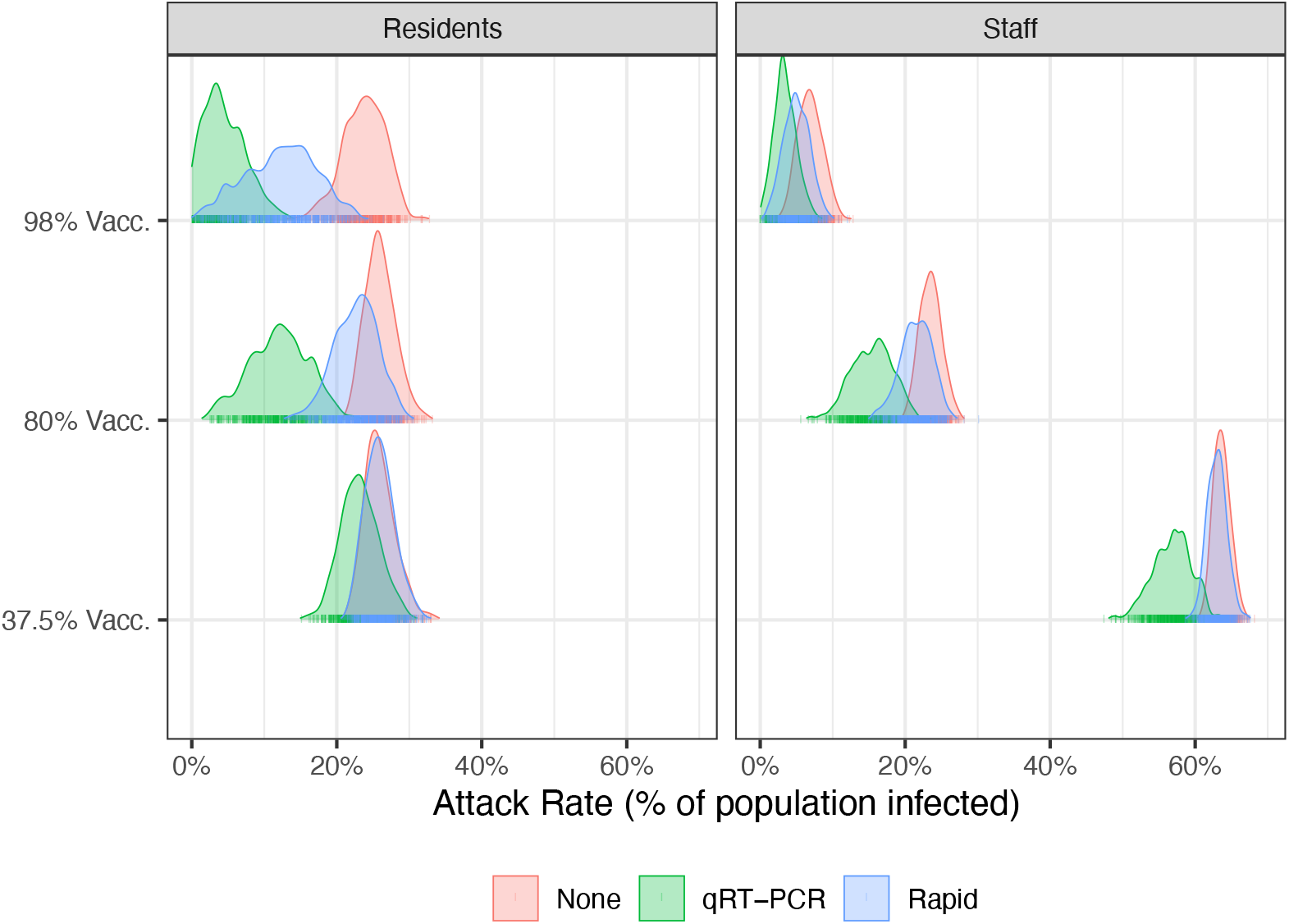
The attack rate in residents (red) and staff (blue) populations over 60-day simulation by vaccination and testing scenario. The distribution represents the variation in results due to stochastic transmission during 500 simulations. Summary statistics are reported in Table S2.

While testing protocols provide protection against transmission, they also lead to staff absenteeism as individuals that test positive are required to isolate. We assume that symptomatic staff will report their illness and self-isolate with some probability. Figure 3 plots the distribution of staff absenteeism measured by the number of days staff are assigned to work but are unable to because of a positive test or self-isolation. Again, higher staff vaccination rates are the most effective mechanism for reducing staff absenteeism because vaccination prevents infection. When comparing the qRT-PCR to the rapid antigen testing protocols, we find that absenteeism is higher under the rapid antigen protocol due to the shorter delay in receiving the test result (immediate versus two days). In the low vaccination scenario (37.5% staff vaccination rate), the absenteeism rate is 85% under the rapid antigen testing protocol versus 80% under the qRT-PCR testing protocol. The difference between the testing protocols becomes smaller as staff vaccination rates increase.

**Figure 3.**
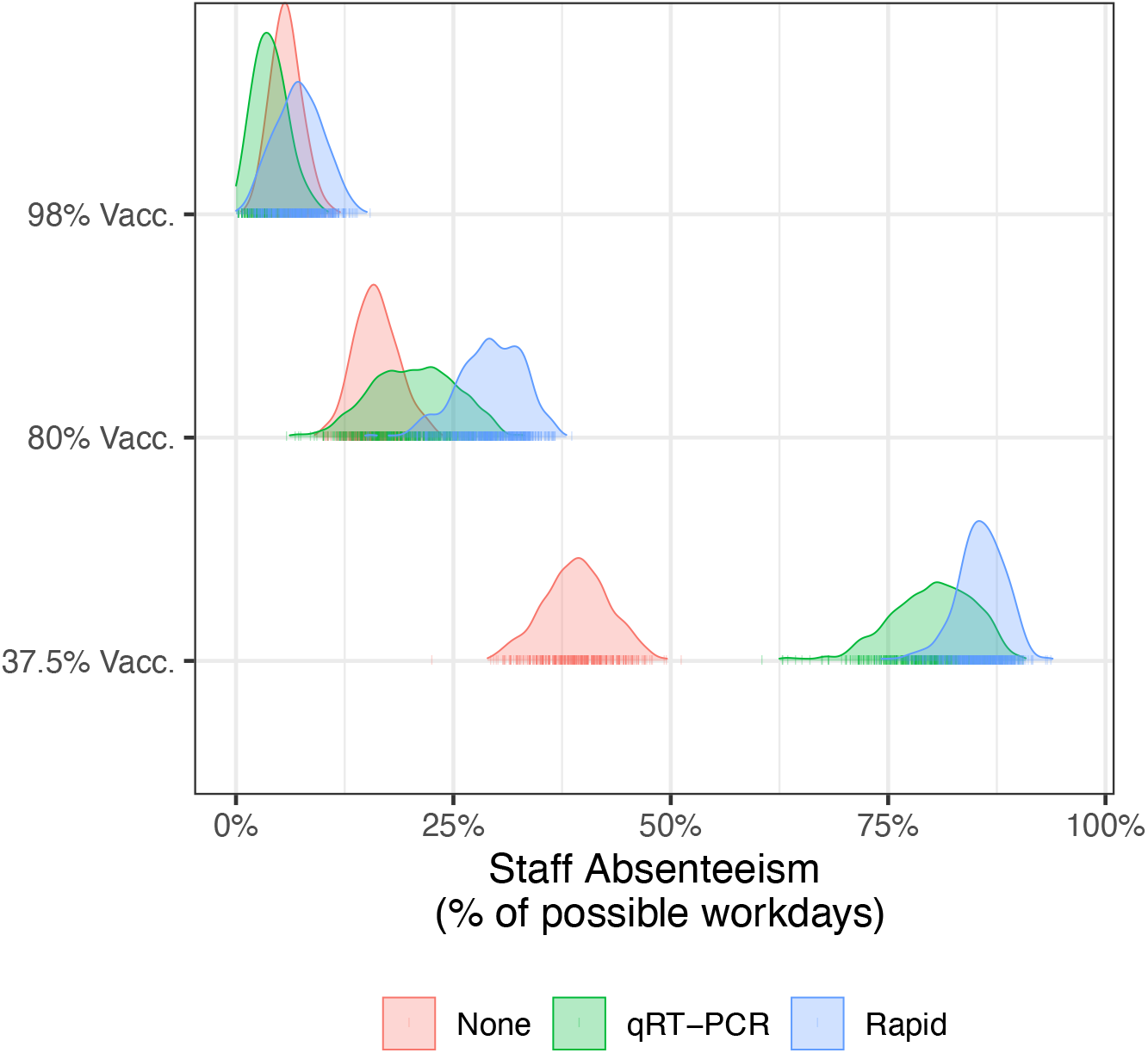
Staff absenteeism by vaccination and testing scenario over 60-day simulation. Absenteeism is defined as workdays missed because of a positive test result or symptomatic self-reporting. The distributions are based on 500 model simulations. Summary statistics are reported in Table S3.

## Discussion

Our model highlights the potential for COVID-19 outbreaks in long-term care facilities even when staff and residents are partially vaccinated and screened regularly. This result is not just theoretical. 627 cases of COVID-19 in staff and residents at 75 skilled nursing facilities were documented from December 28, 2020 to March 31, 2021.^21^ Of those 627, 22 occurred in fully vaccinated individuals more than two weeks after the second dose, and 145 occurred in partially vaccinated individuals.

Despite increasing vaccination and declining cases in Spring 2021, the future of the pandemic is uncertain. There will exist a potential for outbreaks in unvaccinated populations, some of whom may visit family and friends in LTCF. As new variants of the virus emerge, some may pose increased threats or may be more resistant to the existing vaccines. The delta variant is spreading rapidly in Western Colorado and leading to outbreaks in long term care facilities affecting both unvaccinated and vaccinated people.^22^ The duration of immunity from the vaccine is uncertain, leading to speculation about a fall wave and the need for a booster vaccine. Our decision support tool can help LTCF and regional public health departments develop strategies to mitigate the risk of outbreaks and contain them if they emerge.

### Limitations

The results of the LTCF-ABM should be considered along with model limitations. We have tried to use realistic parameters when estimates are available from the literature. However, some parameters are not well studied. We assume a contact structure between staff and residents that is static over the simulation. This may not be true if staff limit their contact with each other and residents in the event of an outbreak. Researches have also documented increased risk to facilities whose staff work at other facilties.^23^ This additional risk is not explicitly modeled here but could be captured by assuming a higher community prevalence quantifying the risk of outside infection to staff.

The testing protocol is determined at the beginning of the simulation and does not change. Facilities that detect a positive case may engage in investigation and more rigorous testing procedures to mitigate the further spread of the virus. However, one important result from our analysis is that when community transmission is present, there is a continual risk that a staff member reintroduces the virus.

## Data Availability

Dashboard code available.

http://www.doi.org/10.5281/zenodo.4984845

## Author Contributions

All authors participated in the concept and design of the project, and preparation of the manuscript, JD led the development of the model.

## Supplement

The corresponding supplemental file contains all simulation parameters and additional detail on the simulation results.

## References

1. Curiskis, A., Kelly, C., Kissane, E. & Oehler, K. Analysis & updates | What We Know—and What We Don’t Know—About the Impact of the Pandemic on Our Most Vulnerable Community. The COVID Tracking Project https://covidtracking.com/analysis-updates/what-we-know-about-the-impact-of-the-pandemic-on-our-most-vulnerable-community (2021).

2. Kim, J. J., Coffey, K. C., Morgan, D. J. & Roghmann, M.-C. Lessons learned - Outbreaks of COVID-19 in nursing homes. Am J Infect Control 48, 1279–1280 (2020).

3. Quinn, C. C., Adams, A. S., Magaziner, J. S. & Gurwitz, J. H. Coronavirus disease 2019 and clinical research in U.S. nursing homes. Journal of the American Geriatrics Society n/a, (2021).

4. Gallichotte, E. N. et al. Longitudinal Surveillance for SARS-CoV-2 Among Staff in Six Colorado Long-Term Care Facilities: Epidemiologic, Virologic and Sequence Analysis. medRxiv 2020.06.08.20125989 (2020) doi:10.1101/2020.06.08.20125989.

5. Gharpure, R. et al. Early COVID-19 First-Dose Vaccination Coverage Among Residents and Staff Members of Skilled Nursing Facilities Participating in the Pharmacy Partnership for Long-Term Care Program — United States, December 2020–January 2021. MMWR Morb Mortal Wkly Rep 70, 178–182 (2021).

6. Cavanaugh, A. M. COVID-19 Outbreak Associated with a SARS-CoV-2 R.1 Lineage Variant in a Skilled Nursing Facility After Vaccination Program — Kentucky, March 2021. MMWR Morb Mortal Wkly Rep 70, (2021).

7. White, E. M. et al. Incident SARS-CoV-2 Infection among mRNA-Vaccinated and Unvaccinated Nursing Home Residents. New England Journal of Medicine (2021) doi:10.1056/NEJMc2104849.

8. Salazar, P. D., Link, N., Lamarca, K. & Santillana, M. High coverage COVID-19 mRNA vaccination rapidly controls SARS-CoV-2 transmission in Long-Term Care Facilities. Res Sq rs.3.rs-355257 (2021) doi:10.21203/rs.3.rs-355257/v1.

9. Moore, J., Groves, T., Pilkerton, C. S., Ashcraft, A. M. & Shrader, C. D. Geriatric antibody response to COVID-19. Journal of the American Geriatrics Society n/a, 1–3 (2021).

10. Bonabeau, E. Agent-based modeling: Methods and techniques for simulating human systems. PNAS 99, 7280–7287 (2002).

11. Naimark, D. et al. Simulation-Based Estimation of SARS-CoV-2 Infections Associated With School Closures and Community-Based Nonpharmaceutical Interventions in Ontario, Canada. JAMA Netw Open 4, e213793 (2021).

12. Truszkowska, A. et al. High-Resolution Agent-Based Modeling of COVID-19 Spreading in a Small Town. Advanced Theory and Simulations 4, 2000277 (2021).

13. Hoertel, N. et al. A stochastic agent-based model of the SARS-CoV-2 epidemic in France. Nat Med 26, 1417–1421 (2020).

14. Romero-Brufau, S. et al. Public health impact of delaying second dose of BNT162b2 or mRNA-1273 covid-19 vaccine: simulation agent based modeling study. BMJ 373, 1087 (2021).

15. Holmdahl, I., Kahn, R., Hay, J. A., Buckee, C. O. & Mina, M. J. Estimation of Transmission of COVID-19 in Simulated Nursing Homes With Frequent Testing and Immunity-Based Staffing. JAMA Netw Open 4, e2110071 (2021).

16. Cuevas, E. An agent-based model to evaluate the COVID-19 transmission risks in facilities. Comput Biol Med 121, 103827 (2020).

17. Paulin, E. Nursing Homes Are Requiring Staff COVID-19 Vaccinations. AARP https://www.aarp.org/caregiving/health/info-2021/nursing-homes-covid-vaccine-mandate.html (2021).

18. Dilliott, J. jakedilliott/ltcf_covid_dashboard: First Release. (Zenodo, 2021). doi:10.5281/zenodo.4984845.

19. Gorges, R. J. & Konetzka, R. T. Staffing Levels and COVID-19 Cases and Outbreaks in U.S. Nursing Homes. Journal of the American Geriatrics Society 68, 2462–2466 (2020).

20. Bubar, K. M. et al. Model-informed COVID-19 vaccine prioritization strategies by age and serostatus. Science 371, 916–921 (2021).

21. Teran, R. A. Postvaccination SARS-CoV-2 Infections Among Skilled Nursing Facility Residents and Staff Members — Chicago, Illinois, December 2020–March 2021. MMWR Morb Mortal Wkly Rep 70, (2021).

22. Wingerter, M. CDC investigating delta variant in Mesa County as it gains growing foothold in Colorado – The Denver Post. Denver Post (2021).

23. Chen, M. K., Chevalier, J. A. & Long, E. F. Nursing home staff networks and COVID-19. PNAS 118, (2021).

24. Prince-Guerra, J. L. et al. Evaluation of Abbott BinaxNOW Rapid Antigen Test for SARS-CoV-2 Infection at Two Community-Based Testing Sites - Pima County, Arizona, November 3-17, 2020. MMWR Morb Mortal Wkly Rep 70, 100–105 (2021).

