## Supplement for "Model-based Evaluation of Continued COVID-19 Risk at Long Term Care Facilities"

Table S1: Simulation settings.

| Input | Simulation values | Source |
|---|---|---|
| Facility | | |
| Number of day staff | 125 | Example facility |
| Number of night staff | 45 | Example facility |
| Number of residents | 90 | Example facility |
| Starting conditions | | |
| Symptomatic residents (%) | 0% | - |
| Asymptomatic staff and residents (%) | 2% | - |
| Staff vaccinated or previously infected in last 6 months (%) | 37.5%, 80%, 98% | 5 |
| Residents vaccinated or previously infected in last 6 months (%) | 78% | 5 |
| Testing | | |
| Test result delay (days) | 2 (qRT-PCR), 0 (Rapid) | - |
| Testing cadence (days) | 7 | - |
| Test sensitivity symptomatic (%) | 95% (qRT-PCR), 64.2% (Rapid) | 24 (Rapid) |
| Test sensitivity asymptomatic (%) | 95% (qRT-PCR), 35.8% (Rapid) | |
| Test specificity (%) | 99.5% | |
| Epidemiology (Advanced) | | |
| R0* | 1.4 | - |
| 14 day case rate/100K | 168 | NYT Covid Tracker |
| Probability of self-isolation | 0.2803 | - |

| Simulation (Advanced) | | |
|---|---|---|
| Start date | 2021-05-01 | - |
| Duration (days) | 60 | - |
| Number of runs | 500 | - |

* Given the heavy usage of personal protective equipment in LTCFs, R0 values estimated for other settings, such as schools or the community, are unlikely to hold in LTCFs.  The R0 value was selected by the authors as something reasonable to explore the impacts of other transmission factors.

Table S2. Attack rate by vaccination and testing scenario over 500 simulations. Mean (10th percentile, 90th percentile).

| | Vaccination Rate | None | qRT-PCR | Rapid |
|---|---|---|---|---|
| Residents | 37.5%. | 26% (23%, 29%) | 23% (20%, 27%) | 26% (23%, 29%) |
| | 80% | 26% (23%, 29%) | 12% (7%, 18%) | 23% (19%, 27%) |
| | 98% | 24% (20%, 28%) | 5% (1%, 9%) | 12% (5%, 19%) |
| Staff | 37.5% | 64% (62%, 65%) | 57% (53%, 61%) | 63% (61%, 65%) |
| | 80% | 24% (22%, 25%) | 15% (12%, 19%) | 22% (19%, 24%) |
| | 98% | 7% (5%, 9%) | 3% (2%, 5%) | 5% (3%, 7%) |

Table S3. Staff absenteeism by vaccination and testing scenario over 500 simulations. Mean (10th percentile, 90th percentile).

| Vaccination Rate | None | qRT-PCR | Rapid |
|---|---|---|---|
| 37.5% | 39% (34%, 44%) | 80% (74%, 86%) | 86% (82%, 89%) |
| 80% | 16% (13%, 20%) | 20% (14%, 27%) | 29% (24%, 34%) |
| 98% | 6% (4%, 8%) | 4% (1%, 7%) | 7% (4%, 11%) |

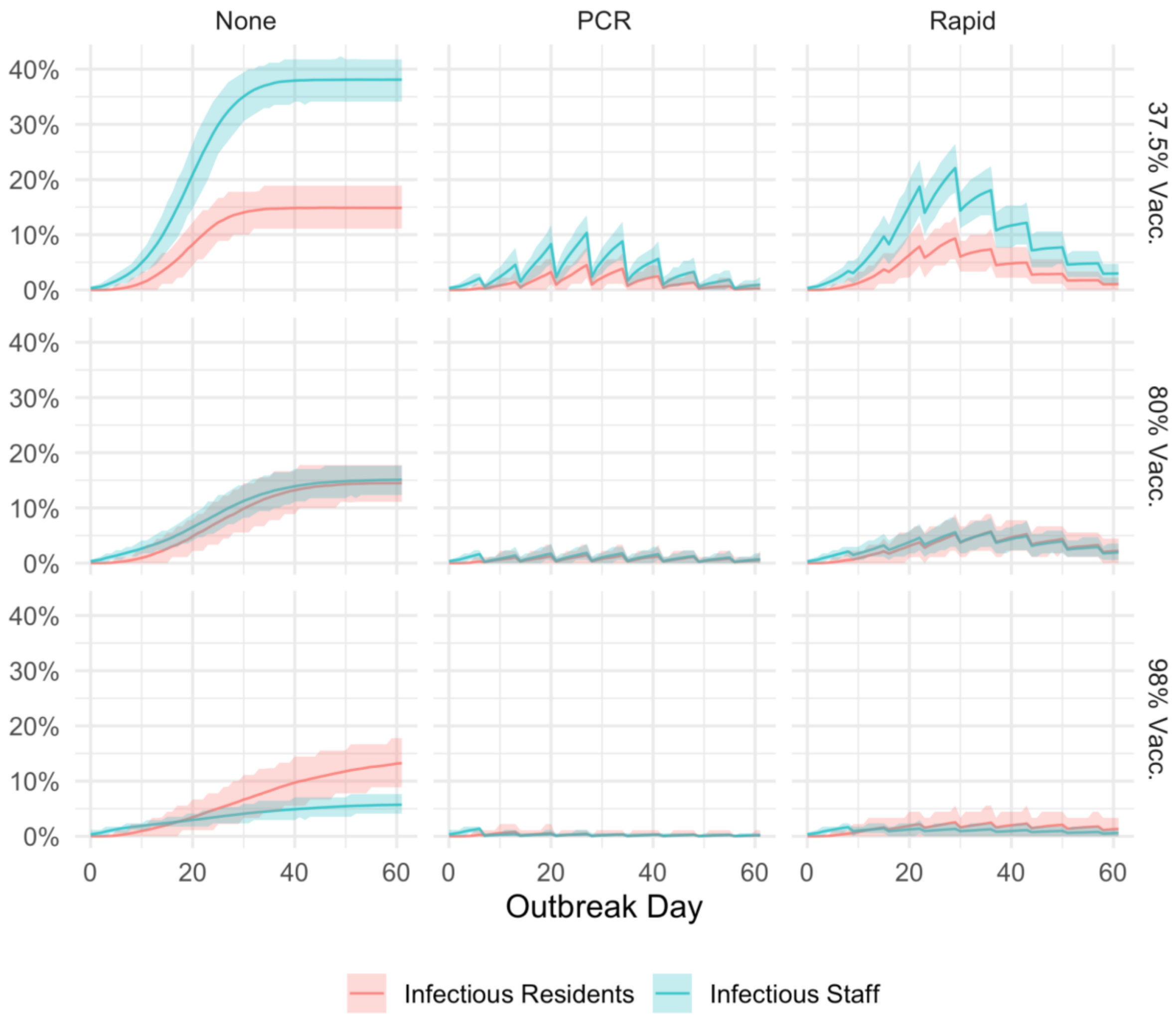


*Figure S1. Number of infectious residents (red) and staff (blue) at the facility by day over 60 days. The line represents the mean daily value and the shaded area represents the tenth (lower) and ninetieth (upper) percentile of the distribution based on 500 model simulations.*
